# Genomic Epidemiology to Investigate the Origins and Zoonotic Implications of Antibiotic-Resistant *Escherichia coli* on Beef and Lamb Meat Sold by Independent Butchers in Wales

**DOI:** 10.64898/2026.03.30.26349758

**Authors:** Jordan E. Sealey, Noora Peltonen, Beatriz Llamazares, Yelyzaveta Moiseienko, Oliver Mounsey, Jasmine Taylor, Laura Wright, Philip Williams, Matthew B. Avison

## Abstract

Many studies have identified antibiotic resistant (ABR) *Escherichia coli* on meat. Appropriate hand hygiene and cooking practices should minimise the risk of gastrointestinal colonisation with ABR *E. coli* found on meat, and the subsequent chance of causing resistant opportunistic extraintestinal infection. There are large gaps in our understanding of the prevalence, origins and zoonotic potential of ABR *E. coli* found on meat, however, and particularly for meat reared in extensive farming systems. Wales is a devolved nation within the United Kingdom having large populations of extensively-reared sheep and beef cattle. To help address knowledge gaps around ABR *E. coli* on extensively reared meat, therefore, beef mince and lamb loin/leg steaks/chops were purchased from 50 (beef) and 46 (lamb) independent butchers across Wales. Following enrichment culture, 200 g meat samples were found to be positive for *E. coli* resistant to amoxicillin (31% positivity), streptomycin (28%), spectinomycin (29%), amoxicillin-clavulanate (11%), 3^rd^ generation cephalosporins (2%) and fluoroquinolones (5%). Phylogenetic analysis confirmed that Welsh lamb meat ABR *E. coli* isolates (n=79) are more closely related to those found in faecal samples collected around sheep (n=352) than around beef cattle (n=361) on Welsh farms. This suggests that faecal contamination at or around slaughter is their primary origin. We found no closely related meat/infection clones (<20 SNPs distant and the same antibiotic resistance genes) when comparing ABR *E. coli* from Welsh meat (n=92) and those causing extraintestinal infections in people (n=2387) in an English region bordering Wales. We conclude, therefore, that the wider zoonotic implications of finding ABR *E coli* on beef and lamb meat sold at independent butchers in Wales are small.

## Introduction

*Escherichia coli* is a focal point for our attempts to mitigate the rising global health threats posed by antibiotic-resistant (ABR) infections. Whilst *E. coli* is an important cause of gastrointestinal infection, in the context of antibiotic resistance its ability to cause opportunistic extraintestinal infections is more troubling. A large proportion of bloodstream infections (BSIs), and most urinary tract infections (UTIs) in humans are cause by extraintestinal *E. coli*. Antibiotic resistance increases morbidity and mortality for both (1–3).

Most opportunistic extraintestinal *E. coli* infections arise due to autoinfection from bacteria carried in the gastrointestinal tract. Hence, colonisation with ABR *E. coli* is associated with ABR extraintestinal infections. Most of these infections in humans are caused by *E. coli* clones from phylogroup B2, which are very rarely found outside of human populations or environments contaminated by human faeces (1,2). As well as commonly carrying acquired antibiotic resistance, such clones, particularly those from sequence types (STs) 131, 1193, 95, and 73, are frequently more virulent, making UTI and associated BSI more likely. However, a moderate proportion of human infections are cause by *E. coli* from phylogroups A, B1 and D, which are more widely distributed and commonly carried in the gastrointestinal tracts of farmed animals (1,2,4–7). There is a concern, therefore, that ingestion of farm-animal-derived ABR *E. coli* might be increasing the burden of ABR extraintestinal infections in humans (2). Whilst evidence for this in real-time transmission/infection terms is very weak (5–7), there has been some interest in the relevance of meat as a vector by which farm-animal-associated ABR *E. coli* might be passed to humans, when appropriate cooking/hand hygiene practices are not well adhered to (8,9). Furthermore, the rise of the trend to feed raw (uncooked) meat to domestic pets adds an additional level of risk for people engage in this, since raw-fed pets are more likely to excrete ABR *E. coli* (10–12).

Many studies have measured the abundance of ABR *E. coli* and other pathogens on meat sold for human consumption. However, very few have considered meat from animals where antibiotic usage is low, for example extensively reared beef (13–19) and lamb/sheep meat (20–22). Such meat might be considered “lower risk” because lower antibiotic usage might be associated with lower rates of resistance. However, this is not well understood. Furthermore, we could find no studies that have attempted to associate ABR *E. coli* found on beef and lamb meat with those found in cattle and sheep faeces. Therefore, it remains possible that any ABR *E. coli* found on meat could be derived from cross-contamination sources, including human sources. It is important to understand these dynamics if we are to suggest interventions that might mitigate potential harm. Finally, most studies of ABR *E. coli* in a One Health context have focused on *E. coli* resistant to critically important antibiotics such as third generation cephalosporin-resistant (3GC-R) or fluoroquinolone-resistant (FQ-R) *E. coli* (6,7,10,20). Since we know these highly resistant populations tend to be clonal (1,6,7,10,20), it is possible that existing studies have given a biased view of ABR *E. coli* transmission dynamics, which have clouded our understanding.

This study was performed as part of a large consortium project aiming to measure and reduce the levels of antibiotic use and resistance on cattle and sheep farms in Wales, a devolved nation within the United Kingdom with a population of 3.2 million people, 9 million sheep/lambs and 1 million beef cattle. Here, we set out to collect much needed data to test hypotheses around the links between ABR *E. coli* present on extensive beef and sheep farms and those found on beef and lamb meat at point-of-sale, and the implications of these bacteria for ABR infections in humans.

## Materials and Methods

### Meat sample collection

We purchased beef and lamb meat from 50 butchers across Wales between April and October 2024. For ease, we have divided Wales into six regions (**Figure S1**). This was a convenience sample, but we attempted to reflect the total number of cattle and sheep in each region by the number of butchers sampled (Table S1). To allow comparisons, we were consistent in purchasing beef mince and lamb leg/loin steaks or chops. At four butchers, an appropriate cut of lamb was not available.

### Bacteriology

Under sterile conditions, 200 g meat was bagged (Stomacher® 400 Classic Standard) alongside 200 mL of *E. coli* Enrichment Broth (EC Broth, Oxoid). Sealed bags were placed into screw-top plastic jars for incubation for 5 h at 37°C and 180 rpm. Enriched culture (500 µL) was mixed with glycerol (25% v/v final), twenty microliters of each mixture was spread onto Tryptone Bile X-Glucuronide agar without antibiotic, or with amoxicillin (8 mg/L), spectinomycin (32 mg/L), streptomycin (32 mg/L), amoxicillin/clavulanate (8/2 mg/L), cefotaxime (2 mg/L), and ciprofloxacin (0.5 mg/L). Plates were incubated at 37°C for 18 h. Up to 10 blue colonies per plate were tested against the same antibiotics to determine resistance phenotype.

### Whole Genome Sequencing (WGS)

One isolate per resistance phenotype per enriched meat sample was selected for WGS, performed by MicrobesNG to achieve a minimum 30-fold coverage. Cells were prepared and genomic DNA purified using standard protocols (MicrobesNG.co.uk). Genomic DNA libraries were prepared using the Nextera XT Library Prep Kit (Illumina, San Diego, USA). Libraries were sequenced on an Illumina NovaSeq 6000 (Illumina, San Diego, USA) using a 250 bp paired end protocol.

### Bioinformatics

Reads were adapter trimmed using Trimmomatic version 0.30 (23) with a sliding window quality cutoff of Q15. De novo assembly used SPAdes version 3.7 (24). Quality control was with QUAST (25). Exclusion criteria were: contigs >500; largest contig <250,000; total genome size <4,500,000 or >6,000,000; GC content <50.00 or >51.00. Contigs were annotated using Prokka 1.11 (26). Fasta files were analysed using ResFinder 4.1, including pointfinder (28), and MLST 2.0 (28). Sequence alignment used Snippy (4.6.0)(https://github.com/tseemann/snippy). SNP distances were determined using SNP-dists (0.8.2) (https://github.com/tseemann/snp-dists). The reference genome was PubMLST reference PDT001296075.1.

### Farm and Human clinical isolates for comparison

We have previously reported the collection of 1874 samples from faecally-contaminated sites on 10 sheep, 12 mixed beef/sheep and 11 beef farms from locations across Wales between April 2022 and December 2024. This survey resulted in the generation of 713 QC-checked whole genome sequences from ABR *E. coli*, based on selection using plates exactly as described above (29).

We also used 2387 sequenced and quality control checked *E. coli* isolates from blood (n=1368) or urine (n=1019) samples deduplicated by patient. They were sequentially collected at Severn Pathology, a diagnostic laboratory serving Bristol, South Gloucestershire, North Somerset, Bath and North East Somerset, and Wiltshire. Bloodstream isolates from 2020 (n=573) and 2023 (n=795) were collected all year round; urinary isolates from 2023 (n=387) were collected for one week in December. These isolates were sequenced irrespective of antibiotic susceptibility phenotype and have already been reported (9,30). Urinary isolates from 2020 (n=336, collected October-December) were resistant to one or more of nitrofurantoin, ciprofloxacin, amoxicillin-clavulanate, or the 3GC cefpodoxime and have been reported previously (31). Urinary Isolates from 2022 (n=296, collected October-December) were resistant to one or more of the following: nitrofurantoin, amoxicillin-clavulanate, or cefpodoxime, and are reported here for the first time.

## Results

### Sample-level positivity for Beef and Lamb meat

Overall, 80/96 (83%) meat samples were positive for *E. coli* following enrichment culture (**Table 1**). Sample-level positivity to European Medicines Agency (EMA) category C/D antibiotics was 31%, 28%, 29% and 11% for amoxicillin, spectinomycin, streptomycin, and amoxicillin-clavulanate-resistant *E. coli*. Positivity for 3GC-R and FQ-R *E. coli* (EMA category B [Restrict]) was 2% and 5%, respectively (**Table 1**).

**Table 1.** Sample-level positivity for *E. coli* and those resistant (-R) to test antibiotics Amoxicillin (Amox), Spectinomycin (Spec), Streptomycin (Strep) and Amoxicillin-Clavulanate (AMC), which are EMA category C/D; Cefotaxime (CTX), a 3GC and ciprofloxacin (CIP), a fluoroquinolone, are EMA category B. The *p*-value of a Fisher’s Exact test comparing positivity in beef and lamb is recorded.

SAMPLE POSITIVITY IN LAMB MEAT
|  | *n* | *E. coli* | Amox-R | Spec-R | Strep-R | CTX-R | CIP-R | AMC-R |
| --- | --- | --- | --- | --- | --- | --- | --- | --- |
|  | 18 | 17 (94%) | 9 (50%) | 10 (56%) | 10 (56%) | 0 | 2 | 5 (28%) |
| North East | 6 | 4 (67%) | 3 (50%) | 3 (50%) | 3 (50%) | 0 | 0 | 2 (33%) |
| Mid West | 4 | 4 (100%) | 2 (50%) | 1 (25%) | 1 (25%) | 0 | 0 | 0 (0%) |
| Mid East | 7 | 6 (86%) | 4 (57%) | 4 (57%) | 4 (57%) | 1 | 1 | 1 (14%) |
| South West | 7 | 7 (100%) | 4 (57%) | 4 (57%) | 4 (57%) | 1 | 0 | 2 (29%) |
| South East | 4 | 4 (100%) | 1 (25%) | 0 (0%) | 1 (25%) | 0 | 1 | 0 (0%) |
| TOTAL | 46 | 42 (91%) | 23 (50%) | 22 (48%) | 23 (50%) | 2 (4%) | 4 (9%) | 10 (22%) |

SAMPLE POSITIVITY IN BEEF MEAT
|  | *n* | *E. coli* | Amox-R | Spec-R | Strep-R | CTX-R | CIP-R | AMC-R |
| --- | --- | --- | --- | --- | --- | --- | --- | --- |
|  | 20 | 14 (70%) | 2 (10%) | 1 (5%) | 2 (10%) | 0 | 1 | 0 |
| North East | 6 | 5 (83%) | 1 (17%) | 1 (17%) | 1 (17%) | 0 | 0 | 0 |
| Mid West | 4 | 3 (75%) | 1 (25%) | 0 (0%) | 1 (25%) | 0 | 0 | 0 |
| Mid East | 7 | 6 (86%) | 2 (29%) | 1 (14%) | 1 (14%) | 0 | 0 | 1 |
| South West | 7 | 5 (71%) | 1 (14%) | 1 (14%) | 0 (0%) | 0 | 0 | 0 |
| South East | 6 | 5 (83%) | 0 (0%) | 1 (17%) | 0 (0%) | 0 | 0 | 0 |
| TOTAL | 50 | 38 (76%) | 7 (14%) | 5 (10%) | 5 (10%) | 0 (0%) | 1 (2%) | 1 (2%) |
| *p Value* |  | **0.06** | **0.0002** | **<0.0001** | **<0.0001** | - | - | **0.003** |

Lamb meat was more commonly positive than beef for *E. coli* resistant to amoxicillin, spectinomycin, streptomycin and amoxicillin-clavulanate (*p*<0.005, Fishers Exact Test; Table 1). Due to low positivity rates, we did not compare rates for *E. coli* resistant to EMA category B agents (ciprofloxacin and cefotaxime).

### WGS analysis of resistance to important antibiotics used in human medicine

In total, 92 ABR *E. coli* isolates (79 from lamb and 13 from beef) were sequenced where data met QC standards and after removing all but the first sequenced isolate from a sample with a particular sequence type (ST) and antibiotic resistance gene (ARG) combination. We found no evidence of virulence genes associated with enteropathogenic *E. coli*.

**Table S1** reports data for the 25/92 sequenced isolates carrying ARGs or mutations associated with resistance to important antibiotics used to treat human *E. coli* infections. Notably, 11/25 isolates came from samples purchased from the same butcher (butcher 5). Both beef and lamb samples from butcher 5 were positive for FQ-R *E. coli* of ST10, ST155 and ST162. The three lamb/beef pairs differed by six single nucleotide polymorphisms (SNPs) (ST162), 49 SNPs (ST155) and zero SNPs (ST10), respectively, and in each case carried identical ARGs, with the last including an *aac*(*3*)*-IId* gentamicin resistance gene (**Table S2**). There were 10 other FQ-R *E. coli* isolates identified across the collection. In one case also carrying *bla*_CTX-M-55_, which confers 3GC resistance and in another also carrying an *mcr1.1* colistin resistance gene. Generally, as expected, FQ resistance was caused by multiple mutations in DNA gyrase and topoisomerase IV genes, *gyrA* and *parC.* One ST428 isolate had only a single mutation in *gyrA*, and carried a mobile quinolone resistance gene, *qnrS1* (**Table S2**). It is known that together with a single *gyrA* mutation, *qnrS1* can confer fluoroquinolone resistance, but is unable to do so alone (32). Additionally, *qnrS1* was found in three fluoroquinolone-susceptible meat isolates (**Table S2**).

The only other 3GC-R *E. coli* from meat was ST969 carrying *bla*_CMY-2_ (**Table S2**). Other isolates carrying ARGs important for human health were two lamb-derived ST69 isolates (each from a different butcher and 27 SNPs apart) carrying gentamicin resistance gene *aac*(*3*)*-VIa*, and two lamb-derived ST847 isolates, each from a different butcher (one SNP apart) carrying fosfomycin resistance gene *fosA7*, (EMA category A [Avoid]). Finally, 30/92 (33%) of all sequenced meat isolates carried a *dfrA* gene, conferring trimethoprim resistance.

### Relationships between lamb meat isolates and those from beef/sheep farms

Among the ABR *E. coli* isolates from lamb meat (79 isolates), 42 STs were represented. Among these, 62 lamb meat isolates were from 27 STs also represented among 362 out of a total of 713 sequenced *E. coli* isolates (**Table S3**) cultured from faecal samples collected on 33 Welsh sheep and/or beef farms (29). The 362 farm isolates from STs shared with lamb meat were more commonly from sheep than beef cattle (Odds Ratio 1.61, 95% confidence intervals 1.12-2.31, p=0.01).

**Table S3** illustrates meat/farm pairs differing by <100 SNPs based on a core-genome alignment of all meat (n=92) and farm (n=713) *E. coli* isolates. For ease of reference, where multiple partners were seen for a meat isolate on the same farm, only the partner with the smallest SNP distance is recorded (**Table S4**). Overall, there were 69 meat/farm pairs (<100 SNP cutoff) divided into 22 separate clones, with 31 meat isolates from 18 butchers having partners from 26 farms.

Of 31 meat isolates paired with farm isolates, 30 were from lamb meat and 82 farm isolates paired with these 30 lamb meat isolates (<100 SNP cutoff; allowing all paired isolates from each farm to count). Lamb meat/farm isolate pairs differing by <100 SNPs were more likely to be pairs involving an isolate from sheep faeces than from beef cattle faeces (Odds Ratio 3.6, 95% confidence intervals 2.2-6.3, p<0.005).

### Geographical separation of related E. coli isolates from meat and farms

The only two meat isolates <100 SNPs different from farm isolates and resistant to important human antibiotics (fosfomycin) are ST847 (**Table S2**; **Table S4**). Although each is from a different butcher, they differ by only 1 SNP and both isolates carry *fosA7*, *strAB*, *bla*_TEM-1_, *tetB* (**Table S2**). These meat isolates are closely related (14-16 SNPs) to two sheep farm isolates – each from a different farm – which also carry *fosA7*, *strAB*, *bla*_TEM-1_, *tetB*.

The locations of the two butchers and two farms, from which these ST847 isolates were recovered, form an approximately 50 x 40 km rectangle. We therefore calculated the geographical distances in a straight line between the sampling locations (butcher or farm) for each isolate in each meat/farm isolate pair (**Table S4**). The geographical distance for each pair was plotted against the SNP distance, but no correlation was observed (R^2^=0.17; **Figure S2**).

**Table S5** lists all pairs of meat isolates differing by <20 SNPs, a cutoff more appropriate to infer possible transmission (33) where each member of the pair is from a different butcher. There were 12 pairs spanning 8 *E. coli* STs across 8 butchers, when plotting the relevant butchers’ shops on a map, they all cluster into a region that is approximately 45 x 50 km (**Figure 1**). The relevant meat samples were all purchased in the same week.

**Figure 1.**
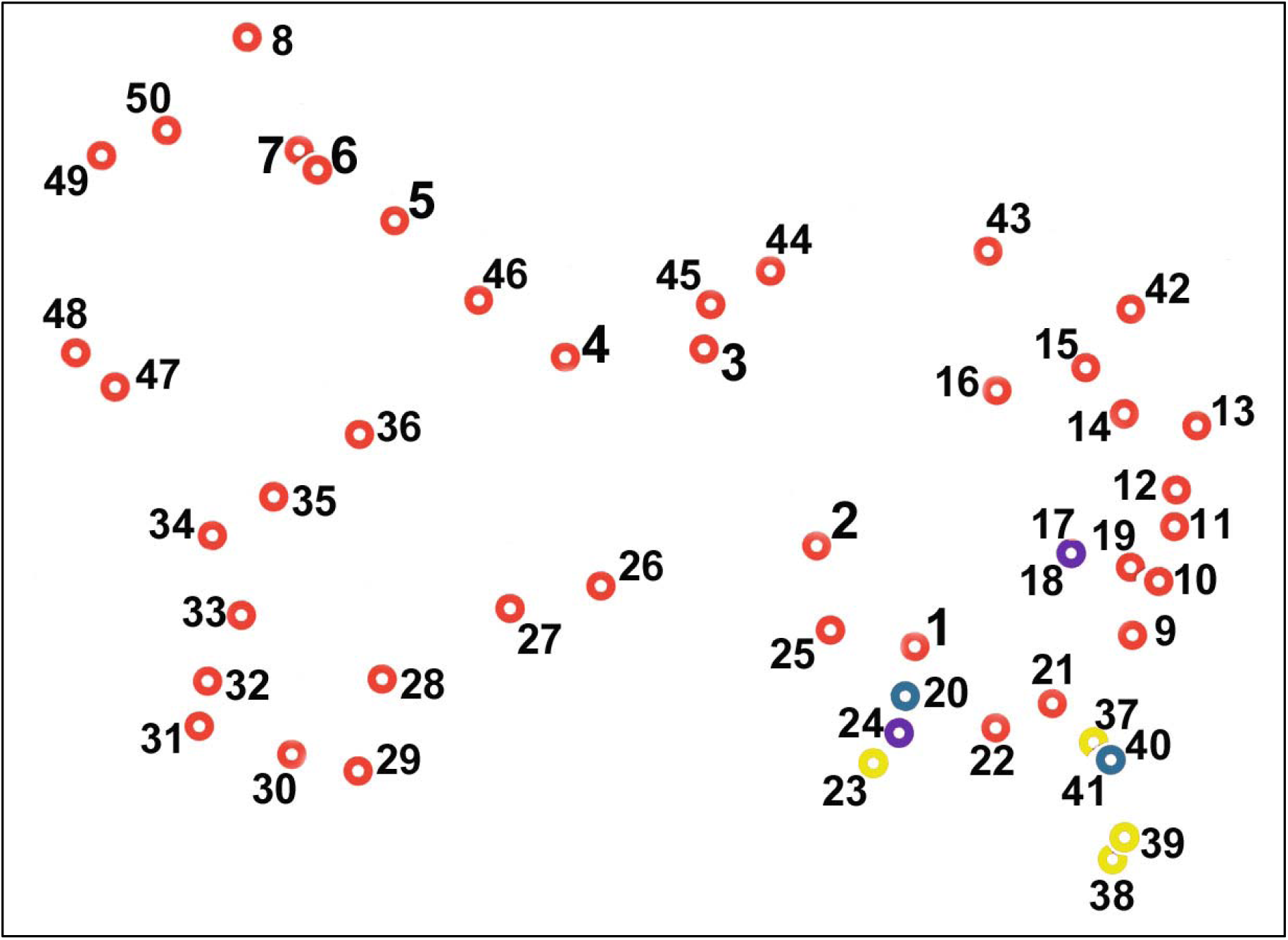
Diagrammatic representation of the locations of the 50 butchers sampled in this study. Red circles represent each, with the numbers used throughout this report. Alternative colours represent clusters of butchers where closely related isolates were found in samples from more than one (colours as in **Table S5**). The 2D distances between each marker accurately reflect the relative geographical locations of each butcher’s shop but the original map used to generate this diagram has undergone multiple transformations to minimize the chance that any shop can be identified.

### Relationships between E. coli isolates from meat and those causing human infections

**Table S5** shows the ST breakdown for 2387 *E. coli* isolates from human UTIs and BSIs in Bristol and surrounding counties – an English region close to the border with Wales – collected between 2020 and 2023, alongside the 92 meat isolates sequenced here. There were 22 STs in common between isolates from lamb meat and human infections, including 54/79 lamb meat isolates and 397/2387 human isolates. This represents a significantly different ST profile (Chi Squared test, *p*<0.00001). There were insufficient beef meat isolates to warrant a separate analysis.

Of 219,604 pairs of isolates compared using core genome alignment, 15 pairs spread across five clones, involved one meat and one human isolate (<100 SNPs cutoff). One clone (comprising two almost identical ST162 meat isolates from butcher 5 and a single urinary isolate) involved SNP distances <20 (**Table 2**). Despite this close relationship (18-19 SNPs) the ST162 meat isolates carried *strAB*, *bla*_TEM-1_, *dfrA14*, *sul2* and *tetA* whilst the urinary isolate carried *aadA1*, *qnrS1*, *bla*_TEM-135_, *dfrA15*, *sul1*, *sul3*, *tetA* and *cmlA1*.

**Table 2.** Pairs of *E. coli* isolates from meat in Wales and those from human infections in Bristol sharing <100 SNPs. The type of meat and infection is noted. Shading indicates <20 SNPs between isolates.

| ST | Meat Isolate | Butcher/ Type (SNP) | Isolate / TYPE (SNP) |  |  |  |  |  |
| --- | --- | --- | --- | --- | --- | --- | --- | --- |
| 428 | 285999 | 23 Lamb | 256820/ BLOOD (96) | 32825/ BLOOD (85) |  |  |  |  |
| 162 | 276352 | 5 Beef | 53881/ URINE (19) |  |  |  |  |  |
|  | 285850 | 5 Lamb (5) | 53881/ URINE (18) |  |  |  |  |  |
| 69 | 286023 | 40 Lamb | 254131/ BLOOD (59) | 255785/ URINE (59) | 256940/ URINE (37) | 267989/ BLOOD (47) | 33310/BLOOD (49) | 40109/ URINE (44) |
| 1722 | 288723 | 40 Lamb | 273993/ BLOOD (33) | 274043/ BLOOD (51) | 39836/ BLOOD (49) |  |  |  |
| 349 | 288839 | 49 Lamb | 255667/ BLOOD (85) | 273965/ BLOOD (95) |  |  |  |  |

## Discussion

To our knowledge this is the first WGS-based analysis of ABR *E. coli* comparing relationships between isolates from beef and lamb meat at point-of-sale and those from extensively reared beef and sheep farms.

Sample-level positivity on meat was high for EMA category C/D antibiotics but low for EMA Category B antibiotics. Whilst it is not appropriate to make direct comparisons with other studies where there are methodological differences, the positivity rates seen here were not unexpected given our previous meat sampling project, at large-chain grocery stores in Bristol, which used an identical methodology (9).

Positivity rates should be considered in the context of the amount of meat tested (200 g) and our use of enrichment, which minimises limitation of detection. In this context, approximately 30% sample positivity for amoxicillin-resistant *E. coli* across all the meat samples is rather low when compared with approximately 60% positivity seen by us among samples collected at faecally-contaminated sites on beef and sheep farms in Wales using a non-enrichment methodology where the limit of detection is 1000-fold higher than that seen for the meat samples here (29).

We noted that lamb meat was considerably more likely to be positive for ABR *E. coli* than beef, which initially surprised us because among our farm samples, ABR *E. coli* positivity is similar for both animal species (29). However, the two types of meat were different: beef mince generally made from steaks (without skin) but lamb steaks/chops with skin present. This skin may be the primary source of *E. coli*, being a surface onto which faecal bacteria might adhere, and future work should investigate this. Related to this point, we report the first evidence that ABR *E. coli* from lamb meat tend to be closely related to those found in faeces collected around sheep. This suggests that faecal contamination at slaughter is important for the transfer of ABR *E. coli* from sheep farms to the consumer via lamb meat.

We also noted close relationships between ABR *E. coli* present on meat being sold at multiple butchers within a small geographical region during the same week. This suggests batches of animals (or carcases) positive for similar ABR bacteria were sold to multiple butchers. One example was our identification of an ST847 fosfomycin-resistant clone found on two farms and in meat sold by two butchers within a small geographical area. However, generally, there was no association between the geographical distance separating the locations of ABR *E. coli* and the phylogenetic distance between them. This is expected, because farmed animals can be transported large distances and dispersed across multiple farms. Hence there can be close relationships between the bacteria carried on multiple farms over long and uncertain distances, as we have seen for beef and sheep farms in Wales (29). Furthermore, transport to sale and slaughter (and of carcases) might also be over long distances.

Despite evidence for a flow of ABR *E. coli* from farms on to meat at point-of-sale, there was no evidence of the ABR *E. coli* present on meat being closely related with those causing resistant human infections. Generally, a 20 SNP distance cut-off is recommended when considering clonality suggestive of transmission (33). Whilst we identified two meat isolates 18 and 19 SNPs different from a human UTI isolate, the ARG profiles of the isolates were different.

Overall, the reported work provides a foundation to our understanding of the sources of ABR *E. coli* on meat sold in Wales and the zoonotic implications thereof. Limitations to our study are mainly due to small scale – 96 meat samples, each a single sample from a single timepoint. Furthermore, the human infection *E. coli* used here were not from Wales, but from Bristol and surrounding counties, though there is considerable movement of people and products between this region and Wales, given their proximity.

## Data Availability

All data produced in the present study are available upon reasonable request to the authors

## Funding

This work was funded by grant 82459 from the Welsh Government Rural Communities - Rural Development Programme 2014-2020 supported by the European Union and the Welsh Government, by grant C003/2023/2024 from the Welsh Government, and by grant MR/T005408/1 from the Medical Research Council.

## Declaration of Competing Interest

This work was conducted as part of the Arwain DGC project which includes farming-related businesses and was funded primarily by the Welsh Government. These organisations had no role in study design, data collection and analysis, decision to publish or preparation of this manuscript.

## Authors’ contributions

MBA, JES designed the study. MBA, PW obtained funding. MBA, JES, OM performed the meat sampling. JES, NP, JT, LW processed meat samples, undertook the bacteriology and prepared isolates for whole genome sequencing. YM, PW collected and prepared human infection isolates for sequencing. NP, BL, JES, OM performed the genomic and statistical analyses. MBA, NP, JES, wrote the manuscript. All authors read and approved the final manuscript.

## References

1. Kaper JB, Nataro JP, Mobley HL. Pathogenic Escherichia coli. Nat Rev Microbiol. 2004;2:123-40. doi: 10.1038/nrmicro818.

2. Jang J, Hur HG, Sadowsky MJ, Byappanahalli MN, Yan T, Ishii S. Environmental Escherichia coli: ecology and public health implications-a review. J Appl Microbiol. 2017;123:570–581. doi: 10.1111/jam.13468.

3. Poirel L, Madec JY, Lupo A, Schink AK, Kieffer N, Nordmann P, Schwarz S. Antimicrobial Resistance in Escherichia coli. Microbiol Spectr. 2018;6:10.1128/microbiolspec.arba-0026-2017. doi: 0.1128/microbiolspec.ARBA-0026-2017.

4. Findlay J, Gould VC, North P, Bowker KE, Williams MO, MacGowan AP, Avison MB. Characterization of cefotaxime-resistant urinary Escherichia coli from primary care in South-West England 2017-18. J Antimicrob Chemother. 2020;75:65–71. doi: 10.1093/jac/dkz397.

5. Mounsey O, Schubert H, Findlay J, Morley K, Puddy EF, Gould VC, North P, Bowker KE, Williams OM, Williams PB, Barrett DC, Cogan TA, Turner KM, MacGowan AP, Reyher KK, Avison MB. Limited phylogenetic overlap between fluoroquinolone-resistant Escherichia coli isolated on dairy farms and those causing bacteriuria in humans living in the same geographical region. J Antimicrob Chemother. 2021;76:3144–3150. doi: 10.1093/jac/dkab310.

6. Ludden C, Raven KE, Jamrozy D, Gouliouris T, Blane B, Coll F, de Goffau M, Naydenova P, Horner C, Hernandez-Garcia J, Wood P, Hadjirin N, Radakovic M, Brown NM, Holmes M, Parkhill J, Peacock SJ. One Health Genomic Surveillance of Escherichia coli Demonstrates Distinct Lineages and Mobile Genetic Elements in Isolates from Humans versus Livestock. mBio. 2019;10:e02693–18. doi: 10.1128/mBio.02693-18.

7. Findlay J, Mounsey O, Lee WWY, Newbold N, Morley K, Schubert H, Gould VC, Cogan TA, Reyher KK, Avison MB. Molecular Epidemiology of Escherichia coli Producing CTX-M and pAmpC β-Lactamases from Dairy Farms Identifies a Dominant Plasmid Encoding CTX-M-32 but No Evidence for Transmission to Humans in the Same Geographical Region. Appl Environ Microbiol. 2020;87:e01842–20. doi: 10.1128/AEM.01842-20.

8. Sealey JE, Astley B, Mounsey O, Avison MB. Poultry-associated nitrofurantoin-resistant and pre-resistant Escherichia coli clones are found in multiple countries and one-health compartments. One Health. 2025;21:101241. doi: 10.1016/j.onehlt.2025.101241.

9. Sealey JE, Astley B, Sealey KL, Daum AM, Kiziltan D, Wright L, Williams P, Avison MB. Similarities Between Antibiotic-Resistant Escherichia coli from Raw Meat, Commercial Raw Dog Food and Those Causing Extraintestinal Human Infections: A Contemporaneous Geographically Focussed Genomic Epidemiology Study. BioRxiv 2026; doi: 10.1101/2024.03.03.583175

10. Mounsey O, Wareham K, Hammond A, Findlay J, Gould VC, Morley K, Cogan TA, Turner KME, Avison MB, Reyher KK. Evidence that faecal carriage of resistant Escherichia coli by 16-week-old dogs in the United Kingdom is associated with raw feeding. One Health. 2022;14:100370. doi: 10.1016/j.onehlt.2022.100370.

11. Sealey JE, Hammond A, Mounsey O, Gould VC, Reyher KK, Avison MB. Molecular ecology and risk factors for third-generation cephalosporin-resistant Escherichia coli carriage by dogs living in urban and nearby rural settings. J Antimicrob Chemother. 2022;77:2399–2405. doi: 10.1093/jac/dkac208.

12. Sealey JE, Hammond A, Reyher KK, Avison MB. One health transmission of fluoroquinolone-resistant Escherichia coli and risk factors for their excretion by dogs living in urban and nearby rural settings. One Health. 2023;17:100640. doi: 10.1016/j.onehlt.2023.100640.

13. Schmidt JW, Vikram A, Doster E, Thomas K, Weinroth MD, Parker J, Hanes A, Geornaras I, Morley PS, Belk KE, Wheeler TL, Arthur TM. Antimicrobial Resistance in U.S. Retail Ground Beef with and without Label Claims Regarding Antibiotic Use. J Food Prot. 2021;84:827–842. doi: 10.4315/JFP-20-376.

14. Bishop H, Evans J, Eze JI, Webster C, Humphry RW, Beattie R, White J, Couper J, Allison L, Brown D, Tongue SC. Bacteriological Survey of Fresh Minced Beef on Sale at Retail Outlets in Scotland in 2019: Three Foodborne Pathogens, Hygiene Process Indicators, and Phenotypic Antimicrobial Resistance. J Food Prot. 2022;85:1370–1379. doi: 10.4315/JFP-22-051.

15. Guragain M, Schmidt JW, Bagi LK, Paoli GC, Kalchayanand N, Bosilevac JM. Antibiotic Resistance and Disinfectant Resistance Among Escherichia coli Isolated During Red Meat Production. J Food Prot. 2024;87:100288. doi: 10.1016/j.jfp.2024.100288.

16. Onwumere-Idolor OS, Kperegbeyi JI, Imonikebe UG, Okoli CE, Ajibo FE, Njoga EO. Epidemiology of multidrug-resistant zoonotic E. coli from beef processing and retail points in Delta State, Nigeria: Public health implications. Prev Vet Med. 2024;224:106132. doi: 10.1016/j.prevetmed.2024.106132.

17. İnat G, Sırıken B, Çiftci A, Erol İ, Başkan C, Yıldırım T. Molecular characterization of extended-spectrum β-lactamases-producing Enterobacteriaceae species in ground beef and chicken meat. Int J Food Microbiol. 2023;398:110228. doi: 10.1016/j.ijfoodmicro.2023.110228.

18. Worku W, Desta M, Menjetta T. High prevalence and antimicrobial susceptibility pattern of salmonella species and extended-spectrum β-lactamase producing Escherichia coli from raw cattle meat at butcher houses in Hawassa city, Sidama regional state, Ethiopia. PLoS One. 2022;17:e0262308. doi: 10.1371/journal.pone.0262308.

19. Gozi KS, Deus Ajude LPT, Barroso MDV, Silva CRD, Peiró JR, Mendes LCN, Nogueira MCL, Casella T. Potentially Pathogenic Multidrug-Resistant Escherichia coli in Lamb Meat. Microb Drug Resist. 2021;27:1071–1078. doi: 10.1089/mdr.2020.0488.

20. Guo S, Aung KT, Leekitcharoenphon P, Tay MYF, Seow KLG, Zhong Y, Ng LC, Aarestrup FM, Schlundt J. Prevalence and genomic analysis of ESBL-producing Escherichia coli in retail raw meats in Singapore. J Antimicrob Chemother. 2021;76:601–605. doi: 10.1093/jac/dkaa461.

21. Abdallah HM, Al Naiemi N, Elsohaby I, Mahmoud AFA, Salem GA, Vandenbroucke-Grauls CMJE. Prevalence of extended-spectrum β-lactamase-producing Enterobacterales in retail sheep meat from Zagazig city, Egypt. BMC Vet Res. 2022 May 20;18(1):191. doi: 10.1186/s12917-022-03294-5. PMID: 35596221; PMCID: PMC9121610.

22. Telli AE, Telli N, Biçer Y, Turkal G, Yılmaz T, Uçar G. Co-Occurrence and Molecular Characterization of ESBL-Producing and Colistin-Resistant Escherichia coli Isolates from Retail Raw Meat. Foods. 2025;14:3573. doi: 10.3390/foods14203573.

23. Bolger AM, Lohse M, Usadel B. Trimmomatic: A flexible trimmer for Illumina Sequence Data. Bioinformatics. 2014;btu170. doi: 10.1093/bioinformatics/btu170.

24. Bankevich A, Nurk S, Antipov D, Gurevich AA, Dvorkin M, Kulikov AS, Lesin VM, Nikolenko SI, Pham S, Prjibelski AD, Pyshkin AV, Sirotkin AV, Vyahhi N, Tesler G, Alekseyev MA, Pevzner PA. SPAdes: A New Genome Assembly Algorithm and Its Applications to Single-Cell Sequencing. J Comput Biol. 2012;19:455–477. doi: 10.1089/cmb.2012.0021.

25. Gurevich A, Saveliev V, Vyahhi N, Tesler G. QUAST: quality assessment tool for genome assemblies. Bioinformatics. 2013;29:1072–5. doi: 10.1093/bioinformatics/btt086.

26. Seemann T. Prokka: rapid prokaryotic genome annotation. Bioinformatics. 2014;30:2068–2069. doi: 10.1093/bioinformatics/btu153.

27. Bortolaia V, Kaas RS, Ruppe E, Roberts MC, Schwarz S, Cattoir V, Philippon A, Allesoe RL, Rebelo AR, Florensa AF, Fagelhauer L, Chakraborty T, Neumann B, Werner G, Bender JK, Stingl K, Nguyen M, Coppens J, Xavier BB, Malhotra-Kumar S, Westh H, Pinholt M, Anjum MF, Duggett NA, Kempf I, Nykäsenoja S, Olkkola S, Wieczorek K, Amaro A, Clemente L, Mossong J, Losch S, Ragimbeau C, Lund O, Aarestrup, F. M. ResFinder 4.0 for predictions of phenotypes from genotypes. J Antimicrob Chemother. 2020;75, 3491–3500. doi: 10.1093/jac/dkaa345.

28. Larsen MV, Cosentino S, Rasmussen S, Friis C, Hasman H, Marvig RL, Jelsbak L, Sicheritz-Pontén T, Ussery DW, Aarestrup FM, Lund O. Multilocus Sequence Typing of Total-Genome-Sequenced Bacteria. J Clin Microbiol. 2012;50,1355–1361. doi: 10.1128/JCM.06094-11.

29. Peltonen N, Sealey JE, Mounsey O, Best C, Llamazares L, Miller W, Moiseienko Y, Sealey KL, Stanton, E, Syvret E, Vass L, Wright L, Reyher KK, Avison MB. Genomic Analyses of Antibiotic-Resistant Escherichia coli From Extensive Beef Cattle and Sheep Farms Identifies Inter-Species and Farm-Farm Sharing as Clonal Dissemination Pathways. bioRxiv. 2025.12.06.692737.

30. Dulyayangkul P, Beavis T, Lee WWY, Ardagh R, Edwards F, Hamilton F, Head I, Heesom KJ, Mounsey O, Murarik M, Pinweha P, Reding C, Satapoomin N, Shaw JM, Takebayashi Y, Tooke CL, Spencer J, Williams PB, Avison MB. Harvesting and amplifying gene cassettes confers cross-resistance to critically important antibiotics. PLoS Pathog. 2024;20:e1012235. doi: 10.1371/journal.ppat.1012235.

31. Dulyayangkul P, Sealey JE, Lee WWY, Satapoomin N, Reding C, Heesom KJ, Williams PB, Avison MB. Improving nitrofurantoin resistance prediction in Escherichia coli from whole-genome sequence by integrating NfsA/B enzyme assays. Antimicrob Agents Chemother. 2024;68:e0024224. doi: 10.1128/aac.00242-24.

32. Wan Nur Ismah WAK, Takebayashi Y, Findlay J, Heesom KJ, Jiménez-Castellanos JC, Zhang J, Graham L, Bowker K, Williams OM, MacGowan AP, Avison MB. Prediction of Fluoroquinolone Susceptibility Directly from Whole-Genome Sequence Data by Using Liquid Chromatography-Tandem Mass Spectrometry To Identify Mutant Genotypes. Antimicrob Agents Chemother. 2018;62:e01814–17. doi: 10.1128/AAC.01814-17

33. Duval A, Opatowski L, Brisse S. Defining genomic epidemiology thresholds for common-source bacterial outbreaks: a modelling study. Lancet Microbe. 2023;4:e349–e357. doi: 10.1016/S2666-5247(22)00380-9.

