## Supplementary Data for "Genomic Epidemiology to Investigate the Origins and Zoonotic Implications of Antibiotic-Resistant *Escherichia coli* on Beef and Lamb Meat Sold by Independent Butchers in Wales"

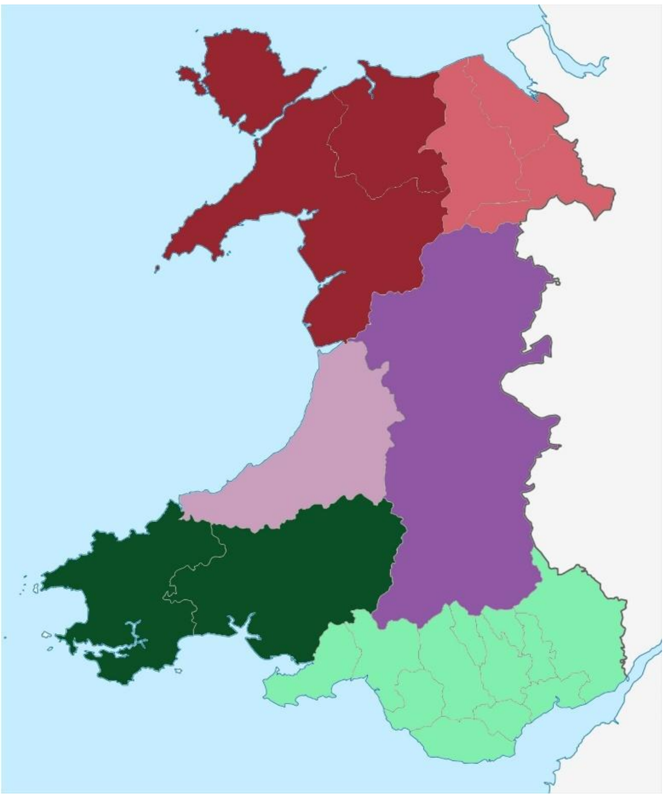

**Figure S1. Map of Principle Areas of Wales grouped into six sampling regions; colour coded as in Table 1**

| | Cattle and Sheep | Butchers | Animals per Butcher | Sampling Farms | Animals per Farm |
|---|---|---|---|---|---|
| North West | 2,581,768 | 20 | 129,088 | 15 | 172,118 |
| North East | 866,054 | 6 | 144,342 | 11 | 78,732 |
| Mid West | 1,002,924 | 4 | 250,731 | 8 | 125,366 |
| Mid East | 4,122,983 | 7 | 588,998 | 6 | 687,164 |
| South West | 1,556,405 | 7 | 222,344 | 12 | 129,700 |
| South East | 1,044,739 | 6 | 174,123 | 18 | 58,041 |
| TOTAL | 11,174,873 | 50 | 223,497 | 70 | 159,641 |

**Table S1. Total number of sheep and cattle (including lambs and calves) recorded for each region, colour coded as in Figure 1. These totals are also divided by the number of butchers' shops visited and Arwain DGC sampling farms in each region.**

| Isolate | Butcher | Meat Type | ST | Fluoroquinolones (B) | 3GCs (B) | Gentamicin (C) | Colistin (B) | Fosfomycin (A) | Trimethoprim (D) | Other EMA Category C/D |
|---|---|---|---|---|---|---|---|---|---|---|
| 285848 | 5 | Lamb | 10* | *gyrA* S83L D87N*; parC* S80I | | *aac(3)-IId* | | | *dfrA17* | *aadA1, aadA2b, strAB, aph(3')-Ia, tetA, cmlA1, sul3* |
| 276351 | 5 | Beef | 10* | *gyrA* S83L D87N*; parC* S80I | | *aac(3)-IId* | | | *dfrA17* | *aadA1, aadA2b, strAB, aph(3')-Ia, tetA, cmlA1, sul3* |
| 285853 | 5 | Lamb | 10 | *gyrA* S83L D87N*; parC* S80I | | | | | *dfrA12* | *aadA2, tetA, tetB, floR, sul1* |
| 285854 | 5 | Lamb | 48 | *qnrS1* | | | | | | *aadA13, strAB, inuG, sul3* |
| 285849 | 5 | Lamb | 155* | *gyrA* S83L D87N*; parC* S80I | | | | | *dfrA14* | *aadA24, strAB, bla*$_{TEM-1}$*, tetA, inuG, sul2* |
| 276354 | 5 | Beef | 155* | *gyrA* S83L D87N*; parC* S80I | | | | | *dfrA14* | *aadA24, strAB, bla*$_{TEM-1}$*, tetA, inuG, sul2* |
| 285850 | 5 | Lamb | 162* | *gyrA* S83L D87N*; parC* S80I | | | | | *dfrA14* | *strAB, bla*$_{TEM-1}$*, tetA, sul2* |
| 276352 | 5 | Beef | 162* | *gyrA* S83L D87N*; parC* S80I | | | | | *dfrA14* | *strAB, bla*$_{TEM-1}$*, tetA, sul2* |
| 285851 | 5 | Lamb | 162 | *gyrA* S83L D87N*; parC* S80I | | | | | *dfrA17* | *aadA5, strAB, bla*$_{TEM-1}$*, tetB, sul2* |
| 285852 | 5 | Lamb | 744 | *gyrA* S83L D87N*; parC* A56T S80I | | | | | *dfrA17* | *aadA5, strAB, aph(3')-Ia bla*$_{TEM-1}$*, tetB, catA1, mphA, sul1, sul2* |
| 285859 | 5 | Lamb | 744 | *gyrA* S83L D87N*; parC* A56T S80I | *bla*$_{CTX-M-55}$ | | | | *dfrA36* | *aadA1, aadA22, aph(3')-Ia, bla*$_{TEM-35}$*, tetB, floR, inuF, sul2, sul3* |
| 285994 | 18 | Lamb | 69* | | | *aac(3)-VIa* | | | *dfrA1* | *aadA1, bla*$_{TEM-1}$*, sul1, sul2* |
| 288712 | 20 | Lamb | 162** | *qnrS1* | | | | | *dfrA14* | *strAB, bla*$_{TEM-1}$*, tetA, sul2* |
| 285998 | 20 | Lamb | 847* | | | | | *fosA7* | | *strAB, bla*$_{TEM-1}$*, tetB* |
| 285981 | 23 | Lamb | 348 | *gyrA* S83L D87N*; parC* S80I | | | | | *dfrA1* | *aadA1, strAB, bla*$_{TEM-1}$*, tetA, sul1, sul2* |
| 285999 | 23 | Lamb | 428 | *gyrA* S83L, *qnrS1* | | | | | | *bla*$_{TEM-1}$*, tetB* |
| 286008 | 33 | Lamb | 969 | | *bla*$_{CMY-2}$ | | | | | |
| 286012 | 34 | Lamb | 69* | | | *aac(3)-VIa* | | | | *aadA1, sul1, sul2* |
| 286016 | 37 | Lamb | 10 | *gyrA* S83L D87N*; parC* S80I | | | *mcr1.1* | | *dfrA14* | *aadA1, aadA2b, strAB, aph(3')-Ia, bla*$_{TEM-1}$*, tetA, cmlA1, floR, mphA, sul2, sul3* |
| 286028 | 37 | Lamb | 10 | *qnrB19* | | | | | | *bla*$_{TEM-135}$*, tetA* |
| 286017 | 37 | Lamb | 162 | *gyrA* S83L D87N*; parC* S80I | | | | | | *tetA, sul2* |
| 288726 | 40 | Lamb | 162** | *qnrS1* | | | | | *dfrA14* | *strAB, bla*$_{TEM-1}$*, tetA* |
| 286034 | 40 | Lamb | 847* | | | | | *fosA7* | | *strAB, bla*$_{TEM-1}$*, tetB* |
| 288840 | 46 | Lamb | 1431 | *gyrA* S83L D87N*; parC* S80I | | | | | *dfrA1, dfrA15* | *bla*$_{TEM-1}$*, tetA* |
| 288816 | 49 | Lamb | 162 | *gyrA* S83L D87N*; parC* S80I | | | | | *dfrA17, sul2* | *aadA5, strAB, bla*$_{TEM-1}$*, tetB* |

**Table S2. Genomic Analysis of *E. coli* resistant to important antibiotics (Ref. 4) used to treat *E. coli* infections in humans (EMA category in brackets) found on meat samples. Shading indicates relatedness (dark <20 SNPs; light 20-49 SNPs) between isolates of the same ST, with stars used to clarify the ST-specific pair being considered.**

| ST | Lamb Meat Isolates | Beef Meat Isolates | Sheep Farm Isolates | Beef Cattle Isolates |
|---|---|---|---|---|
| 10 | 15 | 2 | 33 | 22 |
| 162 | 8 | 1 | 8 | 4 |
| 58 | 4 | | 31 | 31 |
| 69 | 4 | | 13 | 15 |
| 155 | 3 | 1 | 40 | 22 |
| 5908 | 3 | | 1 | |
| 43 | 2 | | 6 | 2 |
| 345 | 2 | | | |
| 744 | 2 | | 5 | 1 |
| 847 | 2 | | 3 | 1 |
| 867 | 2 | | | |
| 2308 | 2 | | 8 | |
| 25 | 1 | | | |
| 48 | 1 | | | |
| 54 | 1 | | 1 | |
| 88 | 1 | | 1 | 4 |
| 93 | 1 | | 3 | |
| 101 | 1 | | 3 | 8 |
| 104 | 1 | | | |
| 154 | 1 | | 3 | 7 |
| 218 | 1 | | | |
| 226 | 1 | | | 2 |
| 348 | 1 | | 1 | 1 |
| 349 | 1 | | 1 | 1 |
| 362 | 1 | | 14 | 9 |
| 428 | 1 | | 1 | |
| 446 | 1 | | 1 | 5 |
| 542 | 1 | | | 1 |
| 969 | 1 | | | |
| 973 | 1 | | 3 | 1 |
| 1086 | 1 | | 13 | 15 |
| 1431 | 1 | | | |
| 1722 | 1 | | 2 | 1 |
| 2556 | 1 | | | |
| 2853 | 1 | | 1 | 1 |
| 5625 | 1 | | | |
| 6115 | 1 | | | |
| 8370 | 1 | | | |
| 9390 | 1 | | | |
| 13897 | 1 | | | |
| 14696 | 1 | 1 | 9 | 3 |
| 7136* | 1 | | | |
| 117 | | 1 | 1 | 1 |
| 295 | | 1 | 1 | |
| 297 | | 1 | 5 | 3 |
| 799 | | 1 | | |
| 1433 | | 1 | 3 | |
| 2179 | | 1 | | |
| 3549 | | 1 | | |
| 17112 | | 1 | | |
| Others | | | 137 | 200 |
| **TOTAL** | **79** | **13** | **352** | **361** |

**Table S3. ST breakdown of sequenced *E. coli* isolates from meat and from Arwain DGC study farms split by meat/animal type**

| Meat Isolate | Butcher/ Type (SNP*) | Farm Isolate(s) /Farm/ TYPE (SNP) | | | | |
|---|---|---|---|---|---|---|
| 285990 | 14 Lamb | 237124/ A/ SHEEP (22) | 276436/ B/ SHEEP (28) | | | |
| 286010 | 33 Lamb | 265948/ C/ SHEEP (59) | 276414/ D/ SHEEP (61) | | | |
| 286014 | 35 Lamb | 237188/ E/ SHEEP (31) | 237207/ F/ SHEEP (36) | 237224/ C/ SHEEP (40) | | |
| 288783 | 44 Lamb (38) | 237188/ E/ SHEEP (37) | 237207/ F/ SHEEP (4) | 237224/ C/ SHEEP (46) | | |
| 286035 | 40 Lamb | 245178/ G/ BEEF (25) | 268025/ H/ BEEF (18) | | | |
| 286024 | 40 Lamb | 276392/ E/ SHEEP (73) | 237195/ I/ SHEEP (78) | 237211/ J/ SHEEP (74) | | |
| 286026 | 26 Lamb | 232129/ K/ SHEEP (11) | | | | |
| 286002 | 23 Lamb | 276395/ L/ SHEEP (42) | | | | |
| 286004 | 28 Lamb (37) | 276395/ L/ SHEEP (25) | | | | |
| 286029 | 38 Lamb (1) | 276395/ L/ SHEEP (41) | | | | |
| 286032 | 39 Lamb (2) | 276395/ L/ SHEEP (42) | | | | |
| 286003 | 24 Lamb | 245123/ M/ BEEF (26) | | | | |
| 286023 | 40 Lamb | 236974/ N/ BEEF (77) | 237114/ O/ SHEEP (25) | 283274/ K/ SHEEP (18) | | |
| 288782 | 42 Lamb | 246920/ B/ SHEEP (4) | | | | |
| 283351 | 3 Lamb | 276343/ P/ BEEF (23) | 276344/ B/ BEEF (15) | | | |
| 286027 | 35 Lamb | 283301/ O/ SHEEP (28) | 246892/ Q/ SHEEP (34) | 246913/ R/ SHEEP (22) | 232395/ S/ SHEEP (22) | 277049/ T/ BEEF (27) |
| 288725 | 40 Lamb | 276407/ A/ SHEEP (90) | 272221/ P/ SHEEP (38) | | | |
| 288839 | 49 Lamb | 283332/ U/ SHEEP (48) | | | | |
| 285845 | 3 Lamb | 277052/ V/ BEEF (23) | 277036/ W/ BEEF (36) | | | |
| 285998 | 20 Lamb | 246904/ T/ SHEEP (16) | 246905/ H/ SHEEP (15) | | | |
| 286034 | 40 Lamb (1) | 246904/ T/ SHEEP (15) | 246905/ H/ SHEEP (14) | | | |
| 285980 | 20 Lamb | 232167/ M/ BEEF (13) | 237203/ D/ SHEEP (29) | 268013/ H/ BEEF (20) | 276400/ O/ SHEEP (22) | 283343/ P/ SHEEP (16) |
| 288723 | 40 Lamb | 237174/ U/ SHEEP (63) | 246968/ F/ SHEEP (35) | | | |
| 285856 | 9 Lamb | 246944/ X/ SHEEP (43) | 246947/ S/ SHEEP (51) | 237218/ Y/ SHEEP (45) | 237202/ D/ SHEEP (44) | 237192/ L/ SHEEP (47) |
| 285996 | 20 Lamb (41) | 246944/ X/ SHEEP (2) | 246947/ S/ SHEEP (10) | 237218/ Y/ SHEEP (4) | 237202/ D/ SHEEP (3) | 237192/ L/ SHEEP (6) |
| 286019 | 37 Lamb | 246933/ P/ SHEEP (12) | | | | |
| 286020 | 38 Lamb (3) | 246933/ P/ SHEEP (11) | | | | |
| 286021 | 39 Lamb (6) | 246933/ P/ SHEEP (6) | | | | |
| 288720 | 23 Lamb | 232425/ Z/ BEEF (37) | 246903/ G/ SHEEP (35) | | | |
| 285983 | 24 Lamb | 283292/ E/ SHEEP (27) | 232146/ P/SHEEP (36) | 276989/ Y/ BEEF (31) | | |
| 286075 | 17 Beef (6) | 283292/ E/ SHEEP (33) | 232146/ P/SHEEP (30) | 276989/ Y/ BEEF (37) | | |

**Table S4. Pairs of *E. coli* isolates from meat and Arwain DGC study farms sharing <100 SNPs. The type of meat/farm is noted. Shading indicates <20 SNPs between isolates. *where isolates from multiple farms are in the same clone all are noted and the SNP distance between each and the first sequenced isolate is recorded in brackets.**

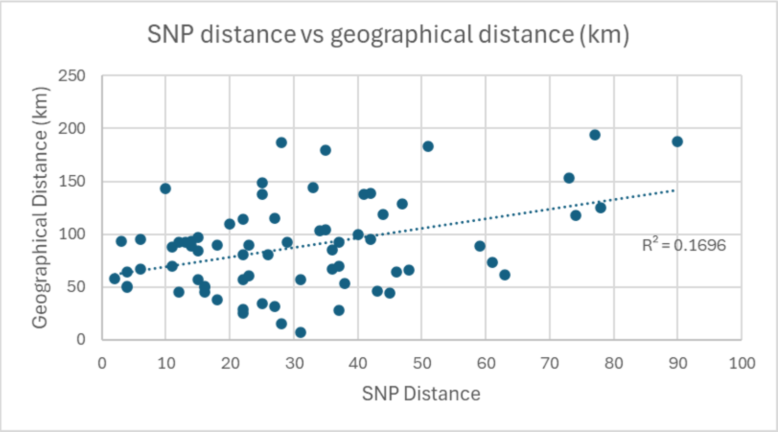


**Figure S2. A plot of SNP distance between each meat/farm isolate pair in Table 5, versus geographical distance between sampling locations.**

| ST | Meat Isolate 1 / Butcher 1 | Meat Isolate 2/ Butcher 2 (SNP) |
|---|---|---|
| 14696 | 286075/ 17 | 285983/ 24 (6) |
| 10 | 285997/ 20 | 286037/ 40 (1) |
| 162 | 288712/ 20 | 288726/ 40 (0) |
| 847 | 288712/ 20 | 286034/ 40 (1) |
| 345 | 285995/ 20 | 288722/ 40 (0) |
| 867 | 286025/ 23 | 286031/ 39 (1) |
| 58 | 286002/ 23 | 286029/ 38 (1) |
| | 286002/ 23 | 286032/ 39 (2) |
| | 286029/ 38 | 286032/ 39 (1) |
| 5908 | 286019/ 37 | 286020/ 38 (3) |
| | 286019/ 37 | 286021/ 39 (6) |
| | 286020/ 38 | 286021/ 39 (9) |

**Table S5. Pairs of *E. coli* isolates from meat from two butchers differing by <20 SNPs. The butcher number is noted. Butchers sharing closely related isolates form three clusters, each differently coloured. The shading colours facilitate locating the butchers' shops in figure 1.**

| ST | Lamb Meat Isolates | Beef Meat Isolates | Human Clinical Isolates |
|---|---|---|---|
| 10 | 15 | 2 | 47 |
| 162 | 8 | 1 | 15 |
| 58 | 4 | | 29 |
| 69 | 4 | | 216 |
| 155 | 3 | 1 | 3 |
| 5908 | 3 | | |
| 744 | 2 | | 4 |
| 2308 | 2 | | |
| 43 | 2 | | |
| 847 | 2 | | 1 |
| 345 | 2 | | 4 |
| 867 | 2 | | |
| 14696 | 1 | 1 | |
| 1086 | 1 | | 1 |
| 362 | 1 | | 7 |
| 973 | 1 | | 1 |
| 93 | 1 | | 9 |
| 101 | 1 | | 4 |
| 154 | 1 | | |
| 446 | 1 | | |
| 88 | 1 | | 22 |
| 348 | 1 | | |
| 1722 | 1 | | 4 |
| 349 | 1 | | 12 |
| 2853 | 1 | | |
| 54 | 1 | | |
| 428 | 1 | | |
| 226 | 1 | | 1 |
| 48 | 1 | | 4 |
| 542 | 1 | | 2 |
| 25 | 1 | | |
| 104 | 1 | | 6 |
| 218 | 1 | | 1 |
| 969 | 1 | | |
| 1431 | 1 | | 4 |
| 2556 | 1 | | |
| 5625 | 1 | | |
| 6115 | 1 | | |
| 8370 | 1 | | |
| 9390 | 1 | | |
| 13897 | 1 | | |
| 7136* | 1 | | |
| 297 | | 1 | 1 |
| 1433 | | 1 | 1 |
| 295 | | 1 | |
| 117 | | 1 | 28 |
| 799 | | 1 | |
| 2179 | | 1 | |
| 3549 | | 1 | |
| 17112 | | 1 | |
| Others | | | 1960 |
| **TOTAL** | **79** | **13** | **2387** |

**Table S6. ST breakdown of sequenced *E. coli* isolates from meat in Wales and from human clinical infections in Bristol**
